# EFFECTS OF MULTIMODAL BALANCE TRAINING WITH AND WITHOUT AUDITORY CUES ON BALANCE, GAIT MOBILITY, RISK OF FALL AND QUALITY OF LIFE IN PATIENTS WITH CHRONIC STROKE

**DOI:** 10.64898/2026.07.20.26357772

**Authors:** Hammad Sattar, Muhammad Hassan Bari, Alishba Mustansar

## Abstract

**Background:** Stroke is a neurological disorder which is defined as the sudden onset of focused or global disruptions in functions of brain caused due to vascular issue which lasts more than 24 hours or sometimes leading to death.

**Objective:** To determine the effects of multimodal balance training with and without auditory cues on balance, gait mobility, risk of fall and quality of life in patients with chronic stroke.

**Methodology:** This randomized controlled trial, conducted at Islam Teaching Hospital and Idrees Hospital Cant. Sialkot, Pakistan, included 21 stroke survivors per group, 42 in total, (aged 45-70, 1-year post-stroke) using non-probability convenient sampling. Group A received multimodal balance training with auditory cues, while Group B received the same training without cues for 12 weeks. Exclusion criteria included respiratory or orthopedic conditions, cognitive disorders (MMSE < 24), aphasia, non-healing ulcers, or osteoporosis. Outcomes (Berg Balance Scale, Time Up and Go Test, Fall Efficacy Scale-International, Stroke Specific Quality of Life Scale) were assessed at baseline, 6 weeks, and 12 weeks.

**Results:** Group A (with auditory cues) showed statistically significant improvements in balance (Berg Balance Scale: median 21 to 47.5, p < .001), gait mobility (Time Up and Go Test: median 26 to 11 seconds, p < .001), fall risk (Fall Efficacy Scale-International: median 61 to 17, p < .001), and quality of life (Stroke Specific Quality of Life Scale: median 91 to 176.5, p < .001) over 12 weeks, outperforming Group B (without auditory cues) in all measures (p < .001 for balance, gait, and fall risk; p = 0.001 and p < .001 for quality of life at 6 and 12 weeks, respectively).

**Conclusion:** Chronic stroke treatments including multimodal balance training with auditory cues have demonstrated significant advantages over a 12-week therapy session. The results demonstrate significant improvements in balance, gait mobility, risk of fall, and quality of life in chronic stroke survivors.

## Introduction

Acute, localized neurological dysfunction brought on by arterial damage to the central nervous system is the clinical definition of stroke, a multifactorial neurological disorder (1). Still a prominent source of disability, it greatly influences functional mobility and quality of life (2). Relying on caretakers for regular duties, about half of stroke survivors need help with everyday activities (3). The World Health Organisation (WHO) defines stroke as a sudden start of targeted or widespread brain function disturbances brought on by vascular problems, lasting more than 24 hours or resulting in death (4).

Of the strokes, around 87% are ischemic, 10% are intracerebral hemorrhages, and 3% are subarachnoid hemorrhages (5). Appropriate public health measures and lifestyle changes can notably help to prevent up to 80% of strokes (3). Stroke risk factors fall into both non-modifiable and changeable categories. While modifiable variables include hypertension, diabetes, obesity, smoking, alcohol intake, physical inactivity, and cardiovascular diseases (6), non-modifiable factors include age, sex, race/ethnicity, and family history. Still the most important changeable risk factor for stroke is hypertension (7). Furthermore very important in stroke prevention are lifestyle choices like physical exercise and nutrition (8).

After a stroke, mobility and freedom are much compromised by gait and balance problems. Commonly showing impaired motor control, postural instability, gait asymmetry, and higher risk of falls are stroke survivors (9). A main determinant of mobility and functional recovery, balance disorder causes long-term impairment and lower quality of life by means of which other factors are limited. With about 73% of stroke survivors having fall-related problems (1), impaired balance raises the risk of falls. Deficit in proprioception, motor control, and postural corrections leads to imbalance (10). Restoring mobility and lowering fall risk in stroke patients depends mostly on rehabilitation programs including balance and gait training (11).

In rehabilitation, multimodal balance training (MMBT) has been increasingly applied to improve balance, postural control, and functional independence. Targeting neuromuscular coordination, MMBT therapies usually include in weight-shifting activities, trunk stability training, and gait training (12). Recent research imply that by increasing neuromuscular synchronisation and spatiotemporal coordination (13), using auditory cues such rhythmic auditory stimulation (RAS) might help to further improve balance and gait performance. Designed to maximize motor responses, gait kinematics, and balance stability in stroke rehabilitation, RAS is a neurologic music therapy tool with rhythmic auditory feedback (12).Although MMBT and RAS have separately shown advantages for increasing post-stroke mobility, little study has looked at their combined impact in stroke rehabilitation.

The purpose of this investigation is to assess the combined effects of multimodal balance training with and without auditory stimuli on the quality of life, gait mobility, fall risk, and balance of chronic stroke survivors. This work aims to give empirical data for an original method of stroke rehabilitation by evaluating the effect of including audio signals into multimodal balance training. Knowing the synergy between MMBT and RAS might provide a fresh approach to improve neuroplasticity, functional recovery, and general well-being in stroke victims. The results of this study could help to develop rehabilitation strategies and raise patient outcomes in stroke recovery.

## Methodology

This study was a nine-month randomized controlled trial (RCT) that was carried out in the physical therapy departments of Imran Idrees Hospital Cant., Sialkot, and Islam Teaching Hospital. The study sought to assess, in patients with chronic stroke, the impact of multimodal balance training with and without auditory cues on balance, gait mobility, risk of fall, and quality of life. Using non-probability convenient sampling, 42 participants in all 21 in each group were gathered; the sample size was computed using Epi-tool and modified for a 10% attrition rate (14). Included were 45–70 year olds who had a history of at least one year post-stroke, a diagnosis of unilateral stroke, Among the exclusion criteria were aphasia, moderate to severe cognitive abnormalities (MMSE <24), severe heart illnesses, uncontrolled hypertension, non-healing ulcers, osteoporosis, and severe visuospatial deficits (15–19).

Participants were randomly assigned to Group A or Group B using a computer-generated random number sequence, produced through an online randomization tool (www.randomizer.org). Randomization was performed by a member of the research team not involved in treatment delivery or outcome assessment. Group A got multimodal balance training with audio signals given via a Google metronome, while Group B received the same training without auditory cues. Over twelve weeks, both groups attended twenty four sessions, each lasting 45 minutes including breaks for warm-up and cool-down. Among the activities for both groups were trunk stability, weight shifts, visual cue walking, and ball-based diagonal reaching. Group A also underwent rhythmic auditory stimulation (RAS) to increase movement amplitude and speed. At baseline, six weeks, and twelve weeks outcome measures including the Berg Balance Scale, Time Up and Go Test, Fall Efficacy Scale-International, and Stroke-Specific Quality of Life Scale were evaluated. Baseline spasticity was assessed using the Modified Ashworth Scale (MAS), a clinical grading tool (0 to 4) used to quantify resistance to passive muscle stretch in patients with neurological conditions such as stroke

Data analysis was done with SPSS version 21.0. Data were compiled using descriptive statistics mean, standard deviation, frequencies, and percentages. Using the Shapiro-Wilk test, which revealed non-normal distribution, the data’s normalcy was investigated and non-parametric tests were then used. Using the Mann-Whitney U test, between-group comparisons were examined; the Friedman test examined within-group variations. One regarded as statistically significant a p-value of ≤0.05. The Research Ethical Committee granted ethical approval; signed informed permission was acquired from every participant, therefore guaranteeing confidentiality, anonymity, and the opportunity to withdraw at any moment without penalty. Following ethical standards, the study told participants of the possible advantages and lack of hazards connected to the research.

## Results

**Table 1:**
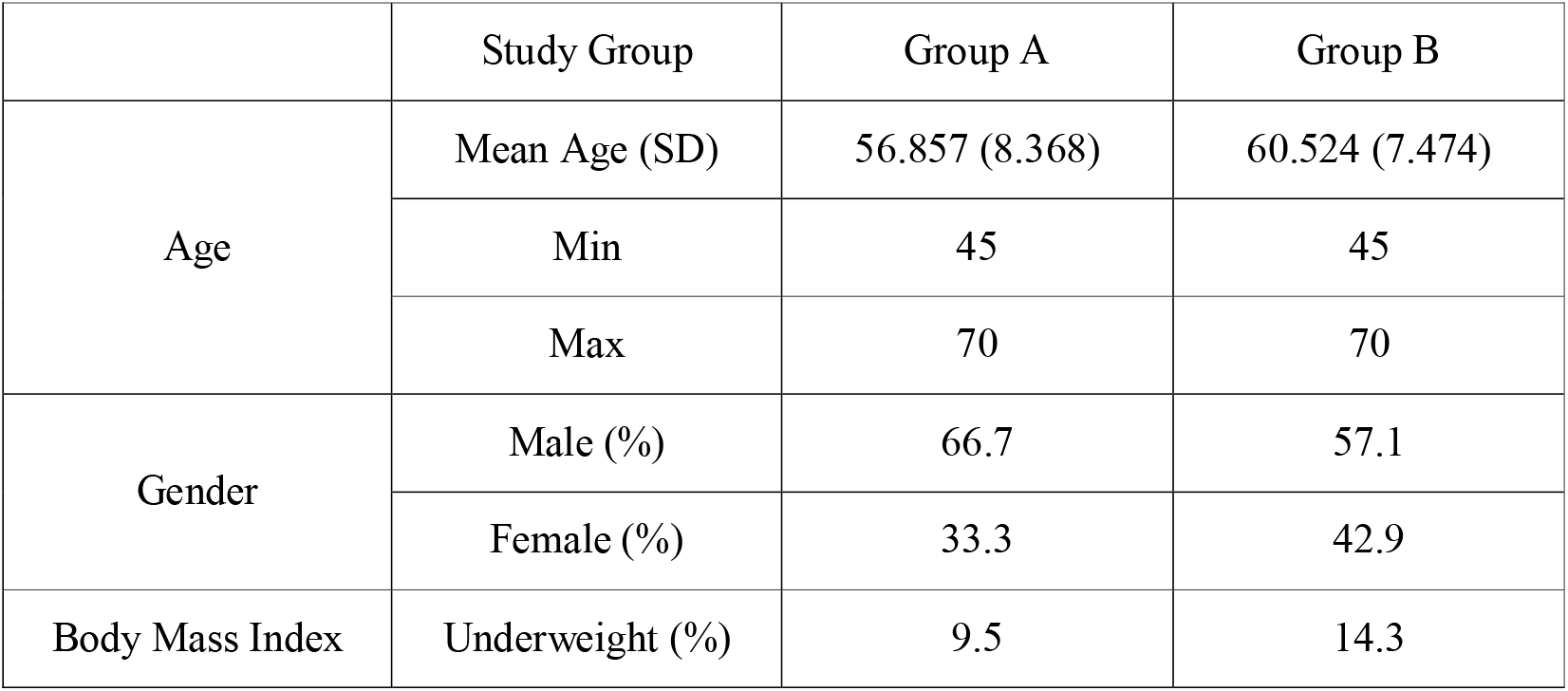
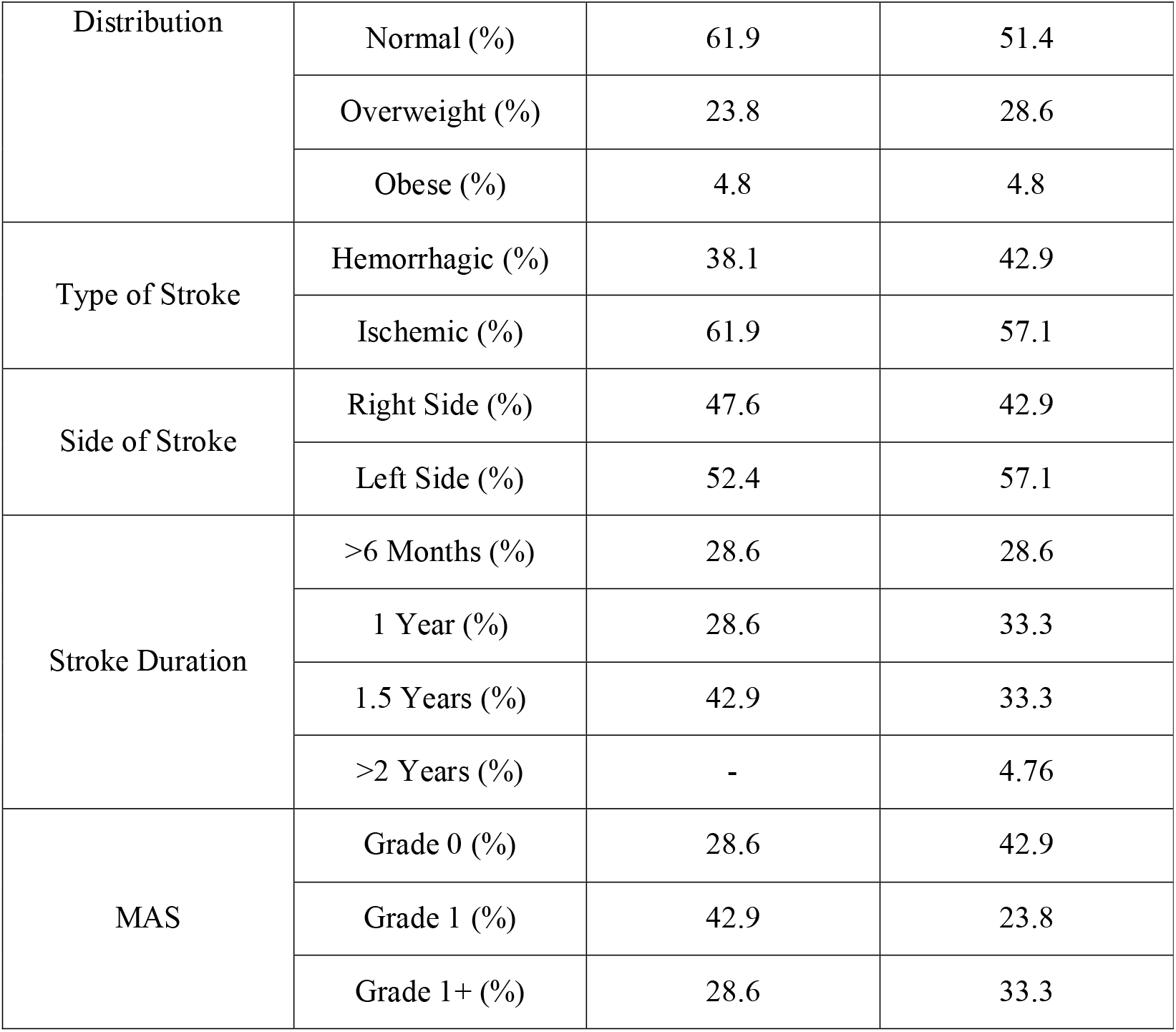
Descriptive Statistics of Demographic and Clinical Characteristics.

Normality of data was assessed using the Shapiro-Wilk test (p < 0.05), which indicated non-normal distribution, consequently, non-parametric tests were employed in the study.

**Table 2:** Inferential Statistics – Within-Group and Between-Group Comparisons (Mann-Whitney U Test with Z Statistic)

| Measure | Timepoint | Group | Median | 25<sup>th</sup> Percentile | 75th Percentile | Z | p-value |
| --- | --- | --- | --- | --- | --- | --- | --- |
| Functional Mobility (TUG) | Baseline | A | 26.0 | 21.5 | 27.0 | 0.816 | 0.415 |
|  |  | B | 23.0 | 20.5 | 26.0 |  |  |
|  | 6 weeks | A | 18.5 | 15.75 | 20.25 | 1.883 | 0.060 |
|  |  | B | 20.0 | 18.0 | 22.5 |  |  |
|  | 12 weeks | A | 11.0 | 8.0 | 18.0 | 2.978 | 0.003 |
|  |  | B | 17.0 | 15.0 | 20.0 |  |  |
| Fall Efficacy Scale | Baseline | A | 60.5 | 54.0 | 64.0 | 1.162 | 0.246 |
|  |  | B | 59.0 | 55.0 | 62.0 |  |  |
|  | 6 weeks | A | 27.0 | 24.0 | 27.0 | 3.317 | 0.001 |
|  |  | B | 27.0 | 25.0 | 29.0 |  |  |
|  | 12 weeks | A | 19.0 | 16.75 | 20.0 | 4.203 | 0.000 |
|  |  | B | 19.0 | 17.0 | 21.0 |  |  |
| Berg Balance Scale | Baseline | A | 21.0 | 21.0 | 24.0 | 1.055 | 0.291 |
|  |  | B | 23.0 | 21.0 | 25.0 |  |  |
|  | 6 weeks | A | 27.0 | 27.0 | 39.0 | 4.433 | 0.000 |
|  |  | B | 27.0 | 24.0 | 29.0 |  |  |
|  | 12 weeks | A | 47.5 | 45.0 | 52.0 | 5.179 | 0.000 |
|  |  | B | 45.0 | 42.0 | 47.0 |  |  |
| Quality of Life Scale | Baseline | A | 91.0 | 87.0 | 95.0 | 1.110 | 0.267 |
|  |  | B | 92.0 | 88.0 | 96.0 |  |  |
|  | 6 weeks | A | 137.0 | 110.0 | 166.25 | 1.851 | 0.064 |
|  |  | B | 110.0 | 105.0 | 140.0 |  |  |
|  | 12 weeks | A | 176.5 | 167.75 | 194.0 | 3.474 | 0.001 |
|  |  | B | 120.0 | 110.0 | 150.0 |  |  |

Note: TUG = Time Up and Go Test. Lower values indicate better performance. Group A significantly outperformed Group B across all measures at 12 weeks ( $p < 0.01$ ).

## Discussion

In comparison to previous studies, the purpose of the current study was to compare the effectiveness of multimodal balance training with and without auditory cues on balance, gait mobility, risk of fall, and quality of life in chronic stroke patients. Over the 12-week intervention period, participants in both groups improved to varying degrees. According to the analysis, Group A showed highly significant improvements, with median scores tracking from baseline to 6 and 12 weeks as follows: TUG improved from 26.0 to 18.5 and 11.0 seconds; FES-I decreased from 60.5 to 27.0 and 19.0; BBS increased from 21.0 to 27.0 and 47.5; and SSQOL increased from 91.0 to 137.0 and 176.5. On the other hand, participants in Group B also showed significant improvements, but to a lesser degree than Group A. Group B’s median scores progressed from baseline to 6 and 12 weeks as follows: TUG shifted from 23.0 to 20.0 and 17.0 seconds; FES-I decreased from 59.0 to 27.0 and 19.0; BBS increased from 23.0 to 27.0 and 45.0; and SSQOL increased from 92.0 to 110.0 and 120.0. Therefore, the analysis demonstrates that while both intervention frameworks improve the status of chronic stroke survivors, multimodal balance training coupled with auditory cues offers a superior therapeutic advantage.

Ambrosini et al. (2020) examined the effects of multimodal balance training on stroke patients’ motor recovery and found significant improvements in balance, gait mobility, and fall risk. The improvements were robust across participants, with statistical analysis showing p-values < 0.05 for both balance and gait mobility. This suggests that multimodal balance training has a significant positive effect on functional recovery. The findings were consistent across various types of stroke and patient severities, emphasizing the efficacy of combining cognitive and physical tasks for optimal recovery (20). In the current study, both groups showed improvements in balance and gait, with Group A (MMBTwAC) demonstrating more significant changes in all measures, including balance and fall efficacy, where the p-values were consistently < 0.01, indicating stronger effects than seen in previous.

Lando et al. (2024) found that multimodal rehabilitation involving physical, proprioceptive, and cognitive training was associated with significant improvements in balance and gait. The study reported p-values of 0.03 for gait speed and 0.02 for balance improvement, supporting the conclusion that multimodal rehabilitation can lead to meaningful gains in physical function. These findings underline the potential for multimodal therapies to target both cognitive and physical recovery processes simultaneously, particularly in individuals with moderate-to-severe stroke disabilities (21). The current study showed that Group A (MMBTwAC) had greater improvements in balance, gait, and fall efficacy, with p-values of 0.01 or lower, suggesting that the addition of auditory cues led to more substantial gains.

Ain et al., 2022 investigated the role of multimodal interventions incorporating sensory feedback such as auditory cues for improving balance and fall prevention. Their study found significant improvements in gait and balance, with p-values of 0.01 for balance and 0.02 for gait stability. This highlighted the potential of multisensory interventions to accelerate motor recovery in stroke patients, with auditory cues showing particular promise in enhancing motor performance (22). The present study corroborates these findings, with Group A (MMBTwAC) showing greater improvements in balance and gait mobility, with p-values consistently < 0.01, suggesting that auditory cues enhance motor coordination more effectively than the standard multimodal balance training.

Pagkalinawan et al. (2021) reported a significant reduction in fall risk for stroke survivors undergoing multimodal rehabilitation, with a p-value of 0.02 for fall efficacy and 0.03 for postural control. Their study emphasized the psychological benefits of multimodal rehabilitation, noting that patients felt more confident in their ability to prevent falls. The results underscore the importance of addressing both the physical and psychological aspects of fall prevention in stroke rehabilitation (23). The current study found a stronger reduction in fall efficacy for Group A (MMBTwAC), with a p-value of 0.01, suggesting that auditory cues provided a greater psychological boost and motor confidence, leading to a more significant reduction in fall risk compared to the non-cue group.

Hlomayi et al. (2023) found that multimodal balance training led to significant improvements in quality of life for stroke patients, with p-values of 0.01 for overall mobility and 0.02 for self-reported well-being. The study suggested that the improvements in physical mobility and fall risk were linked to enhanced psychological and social functioning, which contributed to better quality of life. This research emphasized the holistic benefits of multimodal rehabilitation, including both physical and emotional recovery (24). In the current study, Group A (MMBTwAC) exhibited greater improvements in quality of life, with a p-value of 0.01, suggesting that the added auditory cues not only enhanced physical recovery but also contributed to psychological well-being, leading to more significant gains in quality of life.

## Conclusion

Over a 12-week intervention, this study found that multimodal balance training with auditory signals greatly increases balance, gait mobility, fall risk, and quality of life in people with chronic stroke. The results stress the neuroplastic effects of auditory stimuli, which improve movement patterns and support more efficient functional rehabilitation. These findings highlight how well auditory signals should be used into stroke rehabilitation treatments to maximize recovery results. Emphasizing its function in helping motor learning and enhancing general functional capacities in stroke survivors, the study offers important data supporting the use of rhythmic auditory stimulation as a therapeutic technique.

## Data Availability

All data produced in the present study are available upon reasonable request to the authors

## Abbreviations

MMBT: (Multimodal Balance Training),
MMBTwAC: (Multimodal Balance Training with Auditory Cues, referring to Group A),
RAS: (Rhythmic Auditory Stimulation),
RCT: (Randomized Controlled Trial),
MMSE: (Mini-Mental State Examination),
BBS: (Berg Balance Scale),
TUG: (Time Up and Go Test),
FES**-**I: (Fall Efficacy Scale-International),
SSQOL: (Stroke-Specific Quality of Life Scale),
SPSS: (Statistical Package for the Social Sciences),
SD: (Standard Deviation), and
MAS: (Modified Ashworth Scale).

## LIMITATIONS

- As balance training is a patient dependent training, so during the sessions patients may struggle and difficulty while performing these exercises and also experience anxiety that leads to fear of fall.
- This research involved patients with chronic stroke, so the results do not apply to acute and subacute patients of stroke.
- The use of a single-blind approach, where only the assessors were blinded, introduces potential bias, particularly if participants’ expectations influenced their performance.
- The 12-week intervention with no extended follow-up means the long-term sustainability of the observed improvements in balance, gait, and quality of life could not be assessed.

## RECOMMENDATION

- Future research should include extended follow-up periods to assess the sustainability of improvements in balance, gait mobility, risk of fall and quality of life post-intervention.
- To improve the generalizability of the findings, future studies should involve larger and more diverse populations.
- Combining rhythmic auditory stimulation with advanced rehabilitation technologies, such as virtual reality or wearable sensors, could enhance patient engagement, provide real-time feedback, and optimize rehabilitation outcomes.

## Notes

**Conflict of Interest:** The authors have declared no conflict of interest.

### Competing Interest Statement

The authors have declared no competing interest.

### Clinical Trial

IRCT20240714062437N1

### Author Declarations

Research Ethical Committee of the University of Lahore gave ethical approval for this work.

## References

1. Murphy SJ, Werring DJ. Stroke: causes and clinical features. Medicine. 2020;48(9):561–6.

2. Boddy A, Perry LA, Balasubramanian CK. Effects of a Multi-Modal Gait Training Program in an Individual with Chronic Stroke: a Case Study. Internet Journal of Allied Health Sciences and Practice. 2023;21(4):4.

3. Efremova D, Ciolac D, Zota E, Glavan D, Ciobanu N, Aulitzky W, et al. Dissecting the Spectrum of stroke risk factors in an apparently healthy population: paving the roadmap to primary stroke prevention. Journal of Cardiovascular Development and Disease. 2023;10(2):35.

4. Gao Y, Chen W, Pan Y, Jing J, Wang C, Johnston SC, et al. Dual antiplatelet treatment up to 72 hours after ischemic stroke. New England Journal of Medicine. 2023;389(26):2413–24.

5. Virani SS, Alonso A, Benjamin EJ, Bittencourt MS, Callaway CW, Carson AP, et al. Heart disease and stroke statistics—2020 update: a report from the American Heart Association. Circulation. 2020;141(9):e139–e596.

6. O’Donnell MJ, Chin SL, Rangarajan S, Xavier D, Liu L, Zhang H, et al. Global and regional effects of potentially modifiable risk factors associated with acute stroke in 32 countries (INTERSTROKE): a case-control study. The lancet. 2016;388(10046):761–75.

7. Zhao M, Wang X, He M, Qin X, Tang G, Huo Y, et al. Homocysteine and stroke risk: modifying effect of methylenetetrahydrofolate reductase C677T polymorphism and folic acid intervention. Stroke. 2017;48(5):1183–90.

8. Tu W-J, Wang L-D. China stroke surveillance report 2021. Military Medical Research. 2023;10(1):33.

9. De Luca A, Squeri V, Barone LM, Vernetti Mansin H, Ricci S, Pisu I, et al. Dynamic stability and trunk control improvements following robotic balance and core stability training in chronic stroke survivors: a pilot study. Frontiers in neurology. 2020;11:494.

10. Kang S-M, Kim S-H, Han K-D, Paik N-J, Kim W-S. Physical activity after ischemic stroke and its association with adverse outcomes: A nationwide population-based cohort study. Topics in stroke rehabilitation. 2021;28(3):170–80.

11. Yuen M, Ouyang H, Miller T, Pang MY. Baduanjin qigong improves balance, leg strength, and mobility in individuals with chronic stroke: a randomized controlled study. Neurorehabilitation and neural repair. 2021;35(5):444–56.

12. French B, Thomas LH, Coupe J, McMahon NE, Connell L, Harrison J, et al. Repetitive task training for improving functional ability after stroke. Cochrane database of systematic reviews. 2016(11).

13. Wang L, Peng J-l, Xiang W, Huang Y-j, Chen A-l. Effects of rhythmic auditory stimulation on motor function and balance ability in stroke: a systematic review and meta-analysis of clinical randomized controlled studies. Frontiers in Neuroscience. 2022;16:1043575.

14. In T-s, Jung J-h, Jung K-s, Cho H-y. Effect of sit-to-stand training combined with taping on spasticity, strength, gait speed and quality of life in patients with stroke: A randomized controlled trial. Life. 2021;11(6):511.

15. Buvarp D, Rafsten L, Sunnerhagen KS. Predicting longitudinal progression in functional mobility after stroke: a prospective cohort study. Stroke. 2020;51(7):2179–87.

16. Suzuki K, Matsumaru Y, Takeuchi M, Morimoto M, Kanazawa R, Takayama Y, et al. Effect of mechanical thrombectomy without vs with intravenous thrombolysis on functional outcome among patients with acute ischemic stroke: the SKIP randomized clinical trial. Jama. 2021;325(3):244–53.

17. Pérez-de la Cruz S. Comparison between three therapeutic options for the treatment of balance and gait in stroke: a randomized controlled trial. International journal of environmental research and public health. 2021;18(2):426.

18. Junata M, Cheng KC-C, Man HS, Lai CW-K, Soo YO-Y, Tong RK-Y. Kinect-based rapid movement training to improve balance recovery for stroke fall prevention: a randomized controlled trial. Journal of NeuroEngineering and Rehabilitation. 2021;18:1–12.

19. Ahmed U, Karimi H, Amir S, Ahmed A. Effects of intensive multiplanar trunk training coupled with dual-task exercises on balance, mobility, and fall risk in patients with stroke: a randomized controlled trial. Journal of international medical research. 2021;49(11):03000605211059413.

20. Ambrosini E, Peri E, Nava C, Longoni L, Monticone M, Pedrocchi A, et al. A multimodal training with visual biofeedback in subacute stroke survivors: a randomized controlled trial. European journal of physical and rehabilitation medicine. 2020;56(1):24–33.

21. Lando A, Cacciante L, Mantineo A, Baldan F, Pillastrini P, Turolla A, et al., editors. Multi-Modal versus Uni-Modal Treatment for the Recovery of Lower Limb Motor Function in Patients after Stroke: A Systematic Review with Meta-Analysis. Healthcare; 2024: MDPI.

22. Ain QU, Hassan Z, Ashraf S, Mahjabeen H, Kousar F, Waris M, et al. Comparison Between Effects of Functional Training Program and Conventional Therapy on Postural Control and Functional Mobility in Chronic Stroke. Pakistan Journal of Medical & Health Sciences. 2022;16(02):610–.

23. Pagkalinawan M. Effectiveness of Rhythmic Auditory Stimulation versus Virtual Reality on Gait Velocity and Functional Mobility in Patients with Chronic Stroke: A Meta Analysis: California State University, Fresno; 2021.

24. Hlomayi BY. Effectivity of Combined Rhythmic Auditory Stimulation Rehabilitation Approach on Gait post Stroke. 2023.

